# Stroke volume changes during focal pulsed field vs. radiofrequency ablation for ventricular substrate using Sphere-9 catheter assessed by arterial waveform analysis: a prospective case series

**DOI:** 10.64898/2026.02.23.26346911

**Authors:** Marta Skowronska, Pawel Szymkiewicz, Piotr Gardziejczyk, Ewa Wlazlowska-Struzik, Miron Kusmirek, Jakub Baran

**Affiliations:** Department of Internal Medicine & Cardiology, Medical University of Warsaw, Warsaw, Poland

**Keywords:** pulsed field ablation, lattice tip catheter, stroke volume, ventricular arrhythmia

## Abstract

**Aims:** Catheter ablation using radiofrequency (RF) or pulsed field (PF) energy is an effective treatment method for ventricular arrhythmia (VA). PF offers advantages in lesion formation in anatomically challenging regions. However, its acute effects on ventricular contractility during substrate modification require further elucidation. This study aimed to compare real-time hemodynamic changes associated with PF versus radiofrequency ablation in the left ventricle using stroke volume (SV) as a surrogate for myocardial response in regard to the safety of multiple lesion delivery within scarred myocardium.

**Methods and results:** We conducted a prospective case series study of eight consecutive patients undergoing VA ablation using a dual-energy lattice-tip catheter (Sphere-9, Medtronic). Lesions were delivered to scarred regions identified via intracardiac echocardiography (ICE) and high-resolution 3D mapping. Hemodynamic monitoring was performed using a minimally invasive arterial waveform system (HemoSphere, Edwards Lifesciences). A total of 317 PFA and 41 RF lesions were delivered. PFA applications were associated with a transient SV reduction of 33.1±8.3 ml, with normalization post-delivery. RF lesions resulted in a minimal SV change (≤10% from baseline value). SV reduction following PFA was consistent across lesion locations. All patients achieved post-procedural non-inducibility of clinical VT.

**Conclusion:** PF causes transient but reversible reductions in LV stroke volume during lesion delivery, likely reflecting acute electroporation-induced myocyte stunning rather than irreversible dysfunction. RF lesions did not produce similar changes. These findings suggest a favorable safety profile for PF in ventricular substrate ablation, including in cases of multiple lesion sets, and support its use in regions of scarring. Further studies are warranted to validate these observations and assess long-term outcomes.

## Introduction

Catheter ablation using radiofrequency (RF) or pulsed field (PF) energy is an effective treatment method for ventricular arrhythmia (VA) (1,2). The dependence of PF energy on tissue proximity and not only on rigorous tissue contact has been found useful when delivering lesions to areas characterized by poor stability due to either increased movement or trabeculations (3,4). The lesion dimensions from PF delivery have been consistently reported as deeper and wider than when using RF, further placing PF energy as an increasingly useful alternative or adjunctional treatment to RF delivery (4). However, little is known about the real-time disturbances of ventricular contractility during ablation for VA using PF energy. Such data would be useful to assess the extent of lesions and the location of lesions which may be safely delivered during the index ablation procedure of the ventricular substrate. This study aimed to compare the acute effects of RF and PFA on left ventricular (LV) contractility during ablation for scar-mediated ventricular tachycardia and premature ventricular complexes. We used minimally invasive hemodynamic monitoring to assess stroke volume (SV) changes as a potential surrogate for lesion formation and myocardial response and assess the safety of multiple lesion delivery in scarred tissue.

## Methods

In this preliminary report consisting of a prospectively collected case series, we report on VA ablation performed using a lattice-tip, dual-energy catheter (Sphere-9, Medtronic), and the compatible electroanatomical mapping system (Affera, Medtronic) and its effect on stroke volume. Intracardiac echocardiography (ICE; AcuNav, Siemens), preferably from the left ventricle, was used during all procedures to limit PF application only to scar regions. The ablation target for scar-dependent VT was the identified scarred tissue. The presence of scarring on imaging was defined as hyperechogenicity and narrowing thickness of LV wall in the ICE imaging. High-resolution 3D mapping to assess for low voltage areas was conducted using the ablation catheter by simultaneous recordings from all microelectrodes and automatic annotation as local electrograms from each microelectrode, with the voltage mapping thresholds set according to manufacturer instructions. Lesions were delivered only in areas characterized by both increased hyperechogenicity on ICE and reduced voltage in high-resolution 3D mapping. Lesion delivery was performed using PF, or both PF and RF. Lesion delivery lasted 5.5 seconds for PF and 30 seconds for RF, according to manufacturer instructions. Minimally invasive hemodynamic monitoring was performed using the HemoSphere (Edwards Lifesciences) platform with 5-second averaging for the studied parameters, with the hemodynamic sensor placed in the radial artery. Contractility was assessed by stroke volume (SV). The SV before lesion delivery and immediately post lesion delivery was averaged separately for RF and PF and analyzed, along with the lowest value post lesion delivery for both energies (*ie nadir*). Repetitive lesions of up to three at one site were delivered, consolidation with of PFA with RF was also performed. Lesion formation was controlled for by both the electroanatomical mapping system and newly-formed increased echogenicity in ICE.

## Results

The study population consisted of 8 patients, 2 underwent ablation for PVCs and 6 for VT. All procedures were performed under general anesthesia. The baseline characteristics are presented in Table 1. A total of 317 PF applications and 41 RF applications was delivered, the average number of lesions per patient for RF was 9.1± 4.3 and 70.4 ± 24.0 for PF. The average temporary drop in stroke volume for PF was 33.1±8.3%, whereas RF delivery resulted in an average stroke volume drop below -10% from the baseline value (Table 2). SV normalized to pre-lesion delivery values in all cases in under a minute following lesion delivery. Post-ablation non-inducibility of the clinical VT was achieved in all patients.

**Table 1.** Baseline patient characteristics (n=8) depending on the indication for ablation.

|  | PVC (n=2) | VT (n=6) |
| --- | --- | --- |
| Age (years) | 66 (53-79) | 70 (57-80) |
| Female, n (%) | 1 (33) | 0 |
| ICM, n (%) | 0 | 5 (80) |
| NICM, n (%) | 2 (100) | 1 (12) |
| CKD, n (%) | 0 | 2 (33) |
| LVEF (%) | 37 (22-43) | 29.5 (15-33) |
| LVEDd (mm) | 65.5 (53-78) | 63.5 (53-77) |
| NT-proBNP (pg/ml) | 504 (369-508) | 707.5 (188-4297) |

Data presented median and min-max values or as number followed by percentage.

**Table 2.** Procedural characteristics depending on the indication for ablation.

|  | PVC (n=2) | VT (n=6) |
| --- | --- | --- |
| PFA only | 0 (0) | 2 (33) |
| RFA only | 0 | 0 |
| PF and RF | 2 (100) | 4 (66) |
| Total number of PF applications | 11.5 (7-16) | 40.5 (20-57) |
| Total number of RF applications | 4.5 (2-7) | 4 (0-8) |
| ICE, n(%) | 2 (100) | 6 (100) |
| Steerable sheath, n (%) | 2 (100) | 6 (100) |
| Procedure duration (min) | 112.5 (100-125) | 170 (145-340) |
| Location of lesion delivery and scar | (1) Aorto-mitral continuity<br>(2) Mitral valve annulus | (1) Endocardially apex, basal IVS<br>(2) Endocardially anterior wall and apex<br>(3) Endocardially basal inferolateral wall<br>(4) Epicardially MV<br>(5) Endocardially inferoseptal IVS and RV<br>(6) Endocardially inferolateral wall, inferior wall, mid and basal IVS |
| SV (ml), baseline | 59.5 (48-71) | 65 (40-80) |
| Median drop in SV (ml) | 24.5 (19-30) | 37.5 (24-42) |
| Lowest SV for PFA (ml) | 35 (29-41) | 23.5 (18-29) |
| End SV (ml) | 56.5 (50-63) | 61.5 (45-70) |

Data presented as number followed by percentage or median and min-max values. PVC and VT cases are represented by number in parentheses. Abbreviations: SV- stroke volume.

## Discussion

In this preliminary study evaluating the acute hemodynamic response and safety of multiple PF ablation in scarred left ventricular myocardium, we observed that nearly every PF application resulted in a transient decrease in SV, and all lesions were delivered in areas of scarring. In contrast, RF lesion delivery had a minimal effect on SV.

PFA induces cell death via irreversible electroporation, a nonthermal process that disrupts cell membranes and intracellular homeostasis. In cardiac myocytes, this results in a rapid influx of calcium ions and temporary sarcomere shortening. Prior *in vivo* studies have shown that this effect can persist for several minutes post-ablation, with contractility returning to baseline over time depending on tissue orientation to the electric field (5). Our observed reductions in SV immediately following PFA delivery may therefore represent acute myocyte stunning rather than irreversible myocardial dysfunction.

The variability in the magnitude of SV decline between lesions may be attributed to several factors, including lesion location, fiber orientation relative to the electric field, and anatomical motion. This is consistent with previous findings that PF may be effective in regions where RF lesion delivery is limited (3,4).

Interestingly, we found that RF lesions did not produce similar hemodynamic changes. This aligns with RF’s mechanism of injury — thermal necrosis — which does not acutely impact viable tissue contractility. The absence of a significant SV change following RF delivery may also reflect the smaller number of RF lesions.

The transient nature of SV reduction following PFA — with normalization in all cases — supports the safety of this energy source in ventricular substrate ablation in scarred myocardium. However, as even delivery within scarred myocardium leads to transient contractility dysfunction, it should be carefully applied in viable tissue. We underline the limitations of our monitoring technique: SV was measured using minimally invasive radial artery waveform analysis, which, while practical and real-time, is a surrogate for true contractility and may be influenced by vascular tone or peripheral resistance. More direct measures, such as intracardiac pressure-volume loops would offer more definitive insight.

Finally, the long-term implications of these acute findings remain uncertain. Prior studies have shown that acute PFA lesions undergo significant size regression over time (6). Moreover, while transient decreases in contractility may not pose immediate hemodynamic consequences, their cumulative impact during high-burden lesion sets remains to be investigated.

## Conclusion

In this preliminary study, PF ablation delivered to scarred ventricular myocardium resulted in transient, reversible reductions in stroke volume, with no evidence of sustained contractile dysfunction. These findings support a less sparing use of PF in scarred myocardium. Intra-procedural intracardiac echocardiography remains a valuable adjunct to electroanatomical mapping for identifying arrhythmogenic substrate and guiding lesion delivery during ventricular arrhythmia ablation. These finding warrant larger trials with imaging follow-up.

## Data Availability

All data produced in the present study are available upon reasonable request to the authors

## Reference list

1. Sacher F, Sarkozy A, Pürerfellner H, Steyer A, Lyne J, Coquard C, et al. Safety and efficacy of a lattice-tip catheter for ventricular arrhythmia ablation: the AFFERA Ventricular Arrhythmia Ablation Registry (AVAAR). EP Europace. 2025;27(9):euaf139.

2. Younis A, Tabaja C, Kleve R, Garrott K, Lehn L, Buck E, et al. Comparative efficacy and safety of pulsed field ablation versus radiofrequency ablation of idiopathic LV arrhythmias. JACC Clin Electrophysiol. 2024 Sep;10(9):1998–2009. doi: 10.1016/j.jacep.2024.04.025. Epub 2024 Jun 12. PMID: 38878017.

3. Lozano-Granero C, Hirokami J, Franco E, Tohoku S, Matía-Francés R, Schmidt B, et al. Case series of ventricular tachycardia ablation with pulsed-field ablation: pushing technology further (into the ventricle). JACC Clin Electrophysiol. 2023 Sep;9(9):1990–1994. doi: 10.1016/j.jacep.2023.03.024. Epub 2023 May 24. PMID: 37227358.

4. Kawamura I, Reddy VY, Wang BJ, Dukkipati SR, Chaudhry HW, Santos-Gallego CG, et al. Pulsed field ablation of the porcine ventricle using a focal lattice-tip catheter. Circ Arrhythm Electrophysiol. 2022 Sep;15(9):e011120. doi: 10.1161/CIRCEP.122.011120. Epub 2022 Sep 8. PMID: 36074657; PMCID: PMC9794124.

5. Chaigne S, Sigg DC, Stewart MT, Hocini M, Batista Napotnik T, Miklavčič D, et al. Reversible and irreversible effects of electroporation on contractility and calcium homeostasis in isolated cardiac ventricular myocytes. Circ Arrhythm Electrophysiol. 2022 Nov;15(11):e011131. doi: 10.1161/CIRCEP.122.011131. Epub 2022 Oct 28. PMID: 36306333; PMCID: PMC9665944.

6. Cai C, Ai Z, Wang J, Xu Y, Ju W, Ye C, et al. Pulsed field ablation for idiopathic premature ventricular complexes: evaluation of acute and chronic lesion characteristics with cardiac magnetic resonance imaging. J Interv Card Electrophysiol. 2025 Aug 8. doi: 10.1007/s10840-025-02098-5. Epub ahead of print. PMID: 40779093.

